## Supplementary tables for "The changing epidemiology of human monkeypox – a potential threat? A systematic review"

**S1 Table. Number of Monkeypox Cases by Decade by Country***

| | 1970-1979 | 1980-1989 | 1990-1999<br>Number of Cases | 2000-2009 | 2010-2019 |
|---|---|---|---|---|---|
| Africa | | | | | |
| DRC | 38 | 343 | 511 | 10,027 | 18,788 |
| Nigeria | 3 | — | — | — | 181 |
| Liberia | 4 | — | — | — | 6 |
| Cameroon | 1 | 1 | — | — | 3 |
| Côte d'Ivoire | 1 | 1 | — | — | — |
| Sierra Leone | 1 | — | — | — | 2 |
| Gabon | — | 4 | 9 | — | — |
| Central African Republic | — | 8 | — | — | 61 |
| Congo | — | — | — | 73 | 24 |
| South Sudan | — | — | — | 19 | — |
| Other Continents | | | | | |
| United States | — | — | — | 47 | — |
| United Kingdom | — | — | — | — | 4 |
| Israel | — | — | — | — | 1 |
| Singapore | — | — | — | — | 1 |

* All data reflect the number of confirmed, probable, and/or possible number of monkeypox cases, except for the Democratic Republic of the Congo (DRC) for the years 2000-2009 and 2010-2019, since as of the year 2000, the number of suspected cases was primarily reported by the DRC.

**Table S2. Number of suspected cases versus confirmed, probable, and/or possible cases**

| Author, year (citation) | Study period | Suspected cases (N) | Confirmed, probable, and/or possible cases (n) | Confirmed, probable, and/or possible cases (%) |
|---|---|---|---|---|
| ***Cameroon*** | | | | |
| *WHO, 2018 (67)* | *April - June 2018* | *36 (nr of tested NR)* | *1* | *2.8* |
| *WHO, 2020 (65)* | *Sep 2019* | *1 (nr of tested NR)* | *0* | *0* |
| ***Central African Republic*** | | | | |
| Kalthan, 2018 (33) | Aug - Oct 2016 | 23 of which 7 were tested | 3 | 42.9 |
| *WHO, 2016 (62)* | *Sep - Oct 2016* | *27 (nr of tested NR)* | *3* | *11.1* |
| *WHO, 2017 (60)* | *Feb - April 2017* | *47 (nr of tested NR)* | *5* | *10.6* |
| *WHO, 2017 (61)* | *April-June 2017* | *3 (nr of tested NR)* | *2* | *66.7* |
| *WHO, 2019 (59)* | *March 2018 - June 2019* | *38 (nr of tested NR)* | *25* | *65.8* |
| ***Democratic Republic of the Congo*** | | | | |
| Meyer, 2002 (24) | Feb - Aug 2001 | 31 of which 14 were tested | 7 | 50 |
| Rimoin, 2007 (27) | Jan 2001 - Dec 2004 | 2734 of which 136 were tested | 51 | 37.5 |
| Hoff, 2017 (19) | Nov 2005 - Jan 2008 | 1158 (all were tested) | 785 | 67.8 |
| Nolen, 2016 (25) | July - Dec 2013 | 63 (unclear how many were tested) | 39 | 61.9 |
| McCollum, 2015 (23) | 2011-2014 | 6 (all were tested) | 3 | 50.0 |
| *Laudisoit, 2016 (63)* | *Jan - March 2016* | *160 of which 12 were tested* | *7* | *58.3* |
| *WHO, 2020 (66)* | *Jan - Sep 2020* | *4594 (nr of tested NR)* | *39* | *0.9* |
| ***Liberia*** | | | | |
| *WHO, 2018 (73)* | *Nov 2016 - Dec 2017* | *16 (nr of tested NR)* | *2* | *12.5* |
| ***Nigeria*** | | | | |
| Yinka-Ogunleye, 2019 (44) | Sep 2017 – Sep 2018 | 276 (of which 253 were tested) | 122 | 44.2 |
| ***Republic of the Congo*** | | | | |

| Author, year (citation) | Study period | Suspected cases (N) | Confirmed, probable, and/or possible cases (n) | Confirmed, probable, and/or possible cases (%) |
|---|---|---|---|---|
| Learned, 2005 (46) | April - June 2003 | 12 (all were tested) | 11 | 91.7 |
| Reynolds, 2013 (47) | April - Nov 2010 | 10 of which 4 were tested | 2 | 50.0 |
| Doshi, 2019 (15) | Jan - 5 April 2017 | 43 (unclear how many were tested) | 22 | 51.2 |
| *WHO, 2017 (71)* | *Jan - Sep 2017* | *88 (nr of tested not reported)* | *8* | *9.1* |
| *WHO, 2019 (70)* | *March 2019* | *9 (nr of tested NR)* | *2* | *22.2* |
| ***South Sudan*** | | | | |
| Formenty, 2010 (58) | Sep – Dec 2005 | 49 of which 31 were tested | 19 | 61.3 |
| ***United States*** | | | | |
| CDC, 2003 *(6)* (36-39) | June – July 2003 | 96 (likely all were tested) | 47 | 49.0 |

Note: Citation numbers reflect those that are in the main manuscript text, and those in italics refer to grey literature sources. CDC = United States Centers for Disease Control; nr of tested NR = number of suspected individuals tested is not reported.

**Table S3 Secondary attack rate**

| Author, year (citation) | Secondary attack rate % (n/N) | Comments |
|---|---|---|
| ***Cameroon*** | | |
| Tchokoteu, 1991 (50) | 0 | Denominator of household members was 20, number of neighbors unknown |
| ***Central African Republic*** | | |
| Besombes, 2019 (30) | 0 (0/33) | |
| ***Democratic Republic of the Congo*** | | |
| Breman, 1980 (5) | 7.5 (3/40)<br>3.3 (4/123) | 7.5% refers to very close family members; 3.3% refers to all susceptible contacts. Note that all susceptible contacts possibly include contacts from 9 cases from other countries; data for secondary attack rate not presented per country. |
| Jezek, 1988 (22) | 3 (69/2278) | First generation contacts |
| Jezek, 1986 (20) | 10.2 (4/39) | |
| Aplogan, 1997 (14) | 8 | Editorial note: secondary attack rates were estimated to be 8% (95% CI 5-12%), no further details provided |
| Nolen, 2016 (25) | 50 | Median attack rate in the 16 households: 50% (range 50-100) |
| McCollum, 2015 (23) | 0 (0/30) | Contacts of one case |
| ***Gabon*** | | |
| Meyer, 1991 (54) | 0.3 (1/292) | |
| ***Israel*** | | |
| Erez, 2019 (57) | 0 (0/16) | |
| ***Ivory Coast (Côte d'Ivoire)*** | | |
| Breman, 1977 (51) | 0 | |
| Merouze, 1983 (52) | 0 (0/7) | |
| ***Liberia*** | | |
| Foster, 1972 (42) | 0 (0/44) | |
| | 0 (0/23) | |
| | 0 (0/23) | |
| | 0 (0/136) | |
| ***Sierra Leone*** | | |
| Foster, 1972 (42) | 0 (0/30) | |
| Ye, 2019 (49) | 0 (0/16) | |
| ***Singapore*** | | |
| Yong, 2020 (8) | 0 (0/21) | |
| ***United Kingdom*** | | |
| Vaughan, 2020 (56) | 0.3 (1/288) | |

Note: Citation numbers reflect those that are in the main manuscript text, and those in italics refer to grey literature sources.

### Table S4. Age and sex of confirmed, probable, and/or possible and hospitalised cases from Africa

| Author, year (citation) | Study period | Confirmed cases (n) | Probable cases (n) | Confirmed or probable case (n) | Possible cases (n) | Age median; IQR or range (yrs) | Age case report (yrs) | Age other | Males n (%) |
|---|---|---|---|---|---|---|---|---|---|
| ***Cameroon*** | | | | | | | | | |
| Breman, 1980 (5) | 1979 | 1 | | | | | 3 | | 0 |
| Tchokoteu, 1991 (50) | Dec 1989 | 1 | | | | | 7 | | NR |
| *WHO, 2020 (68)* | *Dec 2019* | *1* | *1* | | | | *1 and NR (mother)* | | *NR* |
| ***Central African Republic*** | | | | | | | | | |
| Herve, 1989 (31) | Likely 1980s | 2 | | | | | 8 and 6 | | 1 (50) |
| Khodakevich, 1985 (34) | Jan 1984 | 6 | | | | | | 5 children; 1 of 22 yrs | 22 year- old is a women. Other NR |
| Berthet, 2011 (29) | June 2010 | 2 | | | | | 14 and 15 | | 2 (100) |
| Nakouné, 2017 (35); Kalthan 2016 (32) | Dec 2015 - Jan 2016 | | | 13 | | 1-41 | | | 7 (53.8) |
| Besombes 2019 (30) | Sep - Oct 2018 | 6 | | | | 4 months – 33 yrs | | | 0 |
| ***Democratic Republic of the Congo*** | | | | | | | | | |
| Breman, 1980 (5) | 1970-1979 | | | 38 | | 7 months – 41 yrs | | | 19 (50) |
| Jezek, 1988 (21) | 1981-1986 | 338 | | | | 4.4 (3 months - 69 yrs) | | | 182 (53.8) |
| Jezek, 1986 (20) | May-July 1983 | 5 | | | | 1-7 | | | 4 (80) |
| Mwanbal, 1997 (13) | Feb 1996 - Feb 1997 | | | | 92 | | | 25 (27.2%) were ≥15 | 51 (55.4) |
| Aplogan, 1997 (14) | Feb 1996 - Oct 1997 | | 304 | | 115 | | | 85% were <16 | NR |
| Meyer, 2002 (24) | Feb - Aug 2001 | 7 | | | | 1-30 | | | 6 (86) |
| Rimoin, 2007 (27) | Jan 2001 - Dec 2004 | 51 | | | | most were <14 | | | 28 (54.9) |
| Rimoin, 2010 (28) | Nov 2005 - Nov 2007 | 760 | | | | 10 (5 days-70 yrs) | | | 472 (62.1) |
| Hoff, 2017 (19) | Nov 2005 - Jan 2008 | 785 | | | | | | 24.2% were <5 | 480 (61.1) |
| Nolen, 2016 (25) | July - Dec 2013 | 20 | 19 | | | 10 (4 months - 68 yrs) | | | 36 (57.1) |
| McCollum, 2015 (23) | 2011-2014 | 3 | | | | | 23-28 | | 2 (66.7) |
| Eltvedt, 2020 (16) | Dec 2016 | | 1 | | | | 4 | | 1 (100) |

| Author, year (citation) | Study period | Confirmed cases (n) | Probable cases (n) | Confirmed or probable case (n) | Possible cases (n) | Age median; IQR or range (yrs) | Age case report (yrs) | Age other | Males n (%) |
|---|---|---|---|---|---|---|---|---|---|
| ***Gabon*** | | | | | | | | | |
| Meyer, 1991 (54) | June 1987 | 1 | 3 | | | 9 months – 9 years | | | 1 (25) |
| No authors, 1992 (53) | Jan, May-July 1991 | 5 | | | | 3-11 | | | NR |
| ***Ivory Coast (Côte d'Ivoire)*** | | | | | | | | | |
| Breman, 1977 (51) | Oct 1972 | | 1 | | | | 5 | | 1 (100) |
| Merouze, 1983 (52) | Jan 1981 | 1 | | | | | 3 | | 0 |
| ***Liberia*** | | | | | | | | | |
| Foster, 1972 (42) | Sep 1970 | 1 | | | | | 4 | | 0 |
| | Sep 1970 | 1 | | | | | 4 | | 1 (100) |
| | Sep 1970 | 1 | | | | | 6 | | 0 |
| | Oct 1970 | 1 | | | | | 9 | | 1 (100) |
| ***Nigeria*** | | | | | | | | | |
| Foster, 1972 (42) | April 1971 | 1 | | | | | 4 | | 0 |
| Breman, 1980 (5) | April 1971 | 1 | | | | | 24 | | 0 |
| Breman, 1980 (5) | Nov 1978 | 1 | | | | | 35 | | 1 |
| Yinka-Ogunleye, 2019 (44) | Sep 2017 - Sep 2018 | 118 | 4 | | | 29; 14 (5 days – 50 yrs) | | | 84 (69) |
| ***Republic of the Congo*** | | | | | | | | | |
| Learned, 2005 (46) | April - June 2003 | 3 | 8 | | | 8 | | | 8 (73) |
| Reynolds, 2013 (47) | April - Nov 2010 | 2 | | | | | 7 and 16 | | 0 |
| Doshi, 2019 (15) | Jan - 5 April 2017 | 7 | 13 | | 2 | 11.5 (1-40) | | | 8 (36.4) |
| ***Sierra Leone*** | | | | | | | | | |
| Foster, 1972 (42) | Dec 1970 | 1 | | | | | 24 | | 1 (100) |
| Reynolds, 2019 (48) | March 2014 | 1 | | | | | 11 months | | 1 (100) |
| Ye, 2019 (49) | March 2017 | 1 | | | | | 35 | | 1 (100) |
| ***South Sudan*** | | | | | | | | | |
| Formenty, 2010 (58) | Sep - Dec 2005 | 10 | 9 | | | 8 months – 32 yrs | | | 9 (47) |

Note: Citation numbers reflect those that are in the main manuscript text, and those in italics refer to grey literature sources. IQR = interquartile range; NR = not reported; yrs = years.

**Table S5. Case fatality rate in confirmed, probable, and/or possible monkeypox cases**

| Author, year (citation) | Study period | Confirmed, probable or possible cases (n) | Case fatality rate n (%) | Age; gender deceased cases |
|---|---|---|---|---|
| ***Cameroon*** | | | | |
| Tchokoteu, 1991 (50) | Dec 1989 | 1 | 0 | |
| *WHO, 2018 (67)* | *April - June 2018* | *1* | *0* | |
| *WHO, 2020 (65)* | *Sep 2019* | *1* | *0* | |
| *WHO, 2020 (68)* | *Dec 2019* | *2* | *1 (50)* | *1; NR* |
| ***Central African Republic*** | | | | |
| Herve, 1989 (31) | Likely 1980s | 2 | 0 | |
| Khodakevich, 1985 (34) | Jan 1984 | 6 | 0 | |
| Berthet, 2011 (29) | June 2010 | 2 | 0 | |
| Nakouné, 2017 (35); Kalthan 2016 (32) | Dec 2015 - Feb 2016 | 13 | 3 (23.1) | 1, 5 and 26; 3M |
| *WHO, 2017 (60)* | *Feb - April 2017* | *5* | *0* | |
| *WHO, 2017 (61)* | *April - June 2017* | *2* | *0* | |
| *WHO, 2019 (59)* | *2 March 2018 - 2 June 2019* | *25* | *3 (12)* | |
| ***Democratic Republic of the Congo*** | | | | |
| Breman, 1980 (5) | 1970-1979 | 38 | 8 (21) | 0-7; 6M, 2F |
| Jezek, 1988 (21) | 1981-1986 | 338 | 33 (9.8) | 0-8; NR |
| Jezek, 1986 (20) | May - July 1983 | 5 | 1 (20) | 1; M |
| Mwanbal, 1997 (13) | Feb 1996 - Feb 1997 | 92 | 3 (3.3) | <3; NR |
| Nolen, 2016 (25) | July - Dec 2013 | 39 | 10 (25.6) | |
| McCollum, 2015 (23) | 2011-2014 | 3 | likely 0 | |
| Eltvedt, 2020 (16) | Dec 2016 | 1 | 1 (100) | 4; NR |
| ***Gabon*** | | | | |
| Meyer, 1991 (54) | June 1987 | 4 | 2 (50) | 0 and 4; 1F, 1M |
| No authors, 1992 (53) | Jan, May-July 1991 | 9 | 0 | |
| ***Israel*** | | | | |

| Author, year (citation) | Study period | Confirmed, probable or possible cases (n) | Case fatality rate n (%) | Age; gender deceased cases |
|---|---|---|---|---|
| Erez, 2019 (57) | Oct 2018 | 1 | 0 | |
| ***Ivory Coast (Côte d'Ivoire)*** | | | | |
| Breman, 1977 (51) | Oct 1972 | 1 | 0 | |
| Merouze, 1983 (52) | Jan 1981 | 1 | 0 | |
| ***Liberia*** | | | | |
| Foster, 1972 (42) | Sep – Oct 1970 | 4 | 0 | |
| *WHO, 2018 (73)* | *Nov 2016 – Dec 2017* | *2* | *0* | |
| ***Nigeria*** | | | | |
| Foster, 1972 (42) | April 1971 | 1 | 0 | |
| Breman, 1980 (5) | April 1971 | 1 | 0 | |
| Breman, 1980 (5) | Nov 1978 | 1 | 0 | |
| *Nigeria Centre for Disease Control (74);* Yinka-Ogunleye 2019 (44) | *2017-2019* | *181** | *9 (5.0)* | mean age 27±14 years in seven cases, including one baby |
| ***Republic of the Congo*** | | | | |
| Learned, 2005 (46) | April – June 2003 | 11 | 1 (9) | 10; F |
| Reynolds, 2013 (47) | April – Nov 2010 | 2 | 0 | |
| Doshi, 2019 (15) | Jan – 5 April 2017 | 22 | 3 (13.6) | 4, 14, 40; 1M, 2F |
| *WHO, 2019 (70)* | *March 2019* | *2* | *0* | |
| ***Sierra Leone*** | | | | |
| Foster, 1972 (42) | Dec 1970 | 1 | 0 | |
| Reynolds, 2019 (48) | March 2014 | 1 | 0 | |
| Ye, 2019 (49) | March 2017 | 1 | 0 | |
| ***Singapore*** | | | | |
| Yong, 2020 (8) | May 2019 | 1 | 0 | |
| ***South Sudan*** | | | | |
| Formenty, 2010 (58) | Sep – Dec 2005 | 19 | 0 | |
| ***United Kingdom*** | | | | |

| Author, year (citation) | Study period | Confirmed, probable or possible cases (n) | Case fatality rate n (%) | Age; gender deceased cases |
|---|---|---|---|---|
| Vaughan, 2018 (55) | Sep 2018 | 2 | 0 | |
| Vaughan, 2020 (56) | Sep 2018 | 1 | 0 | |
| ***United States*** | | | | |
| *Centers for Disease Control and Prevention (6)* | *2003* | *47* | *0* | |

Note: Citation numbers reflect those that are in the main manuscript text, and those in italics refer to grey literature sources. F = female; M = male.

* Two cases diagnosed in the UK were subtracted from the total 183 reported from Nigeria.

**Table S6. Transmission of monkeypox**

| Author, year (citation) | Study period | Number of confirmed, probable and/or possible cases | Mode of transmission | Transmission details |
|---|---|---|---|---|
| ***Cameroon*** | | | | |
| Breman, 1980 (5) | 1979 | 1 | Unknown | Contact with a dead squirrel about two weeks before the illness |
| ***Central African Republic*** | | | | |
| Khodakevich, 1985 (34) | Jan 1984 | 6 | Animal-to-human | A sick animal was eaten. |
| Berthet, 2011 (29) | June 2010 | 2 | Likely animal-to-human | The lesions of both cases developed after hunting and eating a wild rodent |
| Nakouné, 2017 (35) | Dec 2015 – Jan 2016 | 10 | Both | The index case fell sick after killing and cutting up a rodent. Although the authors cannot be 100% certain, the nurses, other members of the family, and different ferryboat drivers were probably contaminated by either the index case or case No. 2 (brother of index case who got sick five days after index case) and not between themselves. |
| Kalthan, 2018 (33) | Aug – Oct 2016 | 3 plus 23 suspected cases | Both | The index case was a hunter and farmer. He had consumed meat that came from the Xerus erythropus species of squirrels, found dead in the forest. All but four patients were secondary cases. |
| Besombes, 2019 (30) | Sep – Oct 2018 | 6 | Both | The index case-patient reported butchering 3 small mammals. The infection of the other 5 family members is considered human-to human transmission. |
| ***Democratic Republic of the Congo*** | | | | |
| Breman, 1980 (5) | 1970-1979 | 38 | Both | Person-to-person spread might have occurred in three cases. In four instances, presumed co-primary cases occurred in the same family. |
| Jezek, 1988 (21) | 1981-1986 | 338 | Both | Animal source: suspected in 245 cases (72.5%); human source in 93 cases (27.5%). |
| Jezek, 1986 (20) | May – July 1983 | 5 | Both | Case 1 ate monkey (rest of the family as well). Circumstances suggest that case 1 infected case 2 and case 2 infected case 3. They belong to one family. Case 4 ate Gambian pouched rat (rest of the family as well). Possibly case 4 was however infected by case 3 by accident in the hospital. Case 5 was likely infected by case 4 (family) |

| Author, year (citation) | Study period | Number of confirmed, probable and/or possible cases | Mode of transmission | Transmission details |
|---|---|---|---|---|
| Aplogan, 1997 (14) | Feb 1996 – Feb 1997 | 419 | Both | Secondary cases: 147 (35%) reported having travelled outside their home village during the 3 weeks preceding disease onset. Of the secondary cases, 53% reported having had antecedent contact with another case-patient in the neighbourhood, 48% in the housing compound, and 42% in an individual household. Primary cases with no apparent association with the clusters in the Akungula/Ekanga occurred in 49 of the 78 affected villages. |
| Meyer, 2002 (24) | Feb - Aug 2001 | 7 | Animal-to-human, and not determined | Outbreak 7: might have been a monkey found dead in the forest that was handled and eaten by the family |
| Nolen, 2016 (25) | Jul – Dec 2013 | 39 | | Nine families showed >1 transmission event, and >6 transmission events occurred within this health zone. |
| McCollum, 2015 (23) | 2011 – 2014 | 3 | Likely all animal-to-human | Case 2: had handled monkeys killed by local hunters, saved monkey meat for his voyage, and then ate this meat over the course of his travel. Case 1 also noted contact with bushmeat before illness onset. |
| ***Gabon*** | | | | |
| Meyer, 1991 (54) | June 1987 | 4 | Unknown | Epidemiological study did not reveal any human transmission evidence, nor the source of infection. |
| No authors, 1992 (53) | Jan, May - July 1991 | 9 | Unknown | Cats, dogs, rats, and mice were found in the homes. In one case a small monkey had been taken into the home a few days before onset of illness. It was not reported whether these could have transmitted MPX. |
| ***Israel*** | | | | |
| Erez, 2019 (57**)** | Oct 2018 | 1 | Likely animal | Case disposed 2 rodent carcasses at his residence. |
| ***Ivory Coast (Côte d'Ivoire)*** | | | | |
| Merouze, 1983 (52) | Jan 1981 | 1 | Unknown | Not possible to prove either animal-human or interhuman contagion |
| ***Liberia*** | | | | |
| Foster, 1972 (42) | Sep 1970 | 1 | Unknown | Playmates with other two cases from Boudua. All three were observed to play with internal organs removed |

| Author, year (citation) | Study period | Number of confirmed, probable and/or possible cases | Mode of transmission | Transmission details |
|---|---|---|---|---|
| | | | | from recently killed monkeys. No evidence of definite monkey contact could be established. No evidence of definite exposure to sick animals, wild or domestic. No exposure to a sick person, either resident or visitor, could be recalled by any of the residents |
| | Sep 1970 | 1 | Unknown | Playmates with other two cases from Boudua. All three were observed to play with internal organs removed from recently killed monkeys. No evidence of definite monkey contact could be established. No evidence of definite exposure to sick animals, wild or domestic. No exposure to a sick person, either resident or visitor, could be recalled by any of the residents |
| | Sep 1970 | 1 | Unknown | Playmates with other two cases from Boudua. All three were observed to play with internal organs removed from recently killed monkeys. No evidence of definite monkey contact could be established. No evidence of definite exposure to sick animals, wild or domestic. No exposure to a sick person, either resident or visitor, could be recalled by any of the residents |
| | Oct 1970 | 1 | Unknown | Occasionally consumed freshly killed monkeys for food. No evidence of definite monkey contact could be established. No evidence of definite exposure to sick animals, wild or domestic. |
| ***Nigeria*** | | | | |
| Foster, 1972 (42) | April 1971 | 1 | Unknown | No evidence of definite exposure to sick animals, wild or domestic, could be obtained. Monkey exposure was highly unlikely. The case had a limited exposure to domestic animals, including dogs, cats, poultry, sheep, goats, and pigs. Residential or farm contact with household rodents and bats was also possible. |
| Breman, 1980 (5) | April 1971 | 1 | Human-to-human | Secondary transmission presumed, mother of other case from Nigeria, April 1971. |

| Author, year (citation) | Study period | Number of confirmed, probable and/or possible cases | Mode of transmission | Transmission details |
|---|---|---|---|---|
| Yinka-Ogunleye, 2019 (44) | Sep 2017 – Sep 2018 | 122 | Both | Of the 122 cases 36 (30%) had an epidemiological link with people with similar lesions before the onset of monkeypox. Of these 36 people, 12 (33%) were epidemiologically linked with a confirmed case. Seven (58%) of these 12 people shared a household or had intimate contact with a confirmed case, four (33%) were inmates in the same prison as a confirmed case, and one was a health worker who treated a confirmed case. Among all confirmed cases, 10 patients reported contact with animals (two with monkeys, two with rodents, two with unspecified wild animal [consumed as meat—ie, bush meat], and four with domestic animals). No one reported contact with sick or dead animals. |
| ***Republic of the Congo*** | | | | |
| Learned, 2005 (46) | April – June 2003 | 11 | Most human-to-human | Case 0 was unavailable for an interview; therefore, the nature of his potential exposures to monkeypox virus is not known. Monkeypox was likely imported with the visit of case 0. Case 1 did own a pet monkey. On visual inspection, the animal appeared to be in good health, and the family could not recall its having been ill within the previous four months. No other potential wild animal exposures were reported for this child, or for the other patients described. The approximate case interval between cases 0 and 1 is consistent with previous descriptions of person-to-person transmission of monkeypox. All subsequent confirmed, probable, and suspect cases in the outbreak had epidemiologic linkage to the Government Hospital in Impfondo, suggesting all further infections were human-to-human. |
| Reynolds, 2013 (47) | April – Nov 2010 | 2 | Unknown | Insufficient evidence. The uncle of the 16-year-old girl and his two sons were reported to have had similar illnesses |

| Author, year (citation) | Study period | Number of confirmed, probable and/or possible cases | Mode of transmission | Transmission details |
|---|---|---|---|---|
| | | | | during the previous month; the uncle died and the two boys had recovered by the time the girl fell ill |
| Doshi, 2019 (15) | Jan – April 2017 | 22 | Both and unknown | Three separate clusters, each in one district: Eyelle: exposure of two cases unknown, 1 likely human-to-human. Dongou: First case was a hunter. For one case exposure was unknown. Six others were hunters but also had contact with cases. The final seven were contacts/family clusters. Impfondo: first case prepared bush meat. Other three were family members. |
| ***Sierra Leone*** | | | | |
| Foster, 1972 (42) | Dec 1970 | 1 | Unknown | No evidence of definite exposure to sick animals, wild or domestic. No evidence of definite monkey contact could be established in the 3 weeks prior to the onset of rash disease. |
| Reynolds, 2019 (48) | March 2014 | 1 | Unknown | The child's mother denied that he had had any contact with persons exhibiting a monkeypox-like illness in the 2 weeks before onset of his illness. The mother also denied that the child had had any history of contact with animals. However, both the mother and father of the boy stated that they regularly prepare and consume meat from wild animals. The mother and father also confirmed that small rodents were sometimes present in the family house. |
| Ye, 2019 (49) | March 2017 | 1 | Animal | The patients had been hunting and eating squirrels ≈10 days before becoming ill, and traveling to Pelewahun gee bu in Bo district 3 days before symptoms began. |
| ***Singapore*** | | | | |
| Yong, 2020 (8**)** | May 2019 | 1 | Likely animal | Ingestion of barbecued bushmeat that might have been contaminated. Patient did not handle raw meat and had no exposure to wild animals or their products, had no contact with rodents or with persons with pox-like illnesses. |
| ***South Sudan*** | | | | |

| Author, year (citation) | Study period | Number of confirmed, probable and/or possible cases | Mode of transmission | Transmission details |
|---|---|---|---|---|
| Formenty, 2010 (58) | Sep – Dec 2005 | 19 | Human-to-human and unknown | Person-to-person transmission was documented in 3 chains associated with the activities of a traditional healer and tooth extractor. Fourteen case-patients reported contact with a suspected monkeypox case-patient before onset of symptoms; 1 case-patient was probably exposed to infected material during his hospitalization at the MSF-F hospital; and 6 case-patients did not report any known likely mode of infection. Among those 6, 3 reported that the rash began appearing around a pre-existing wound, which could have been from the bite of an infected animal. None reported contact with wild animals. |
| ***United Kingdom*** | | | | |
| Vaughan, 2018 (55) | Sep 2018 | 2 | Unknown and likely human-to-human | Case 2: contact with an individual with a monkeypox-like rash at a large family event and consumption of bush meat during his visit to a rural area of Nigeria. |
| Vaughan, 2020 (56) | Sep 2018 | 1 | Human-to-human | The only exposure risk identified during assessment was the changing of potentially contaminated bedding, when case 2 had multiple skin lesions, but before a diagnosis of monkeypox had been considered. |
| ***United States*** | | | | |
| Reynolds, 2007 (41); *CDC (6*) | 2003 | 47 | Animal-to-human | All reported exposure to ill or infected prairie dogs |

Note: Citation numbers reflect those that are in the main manuscript text, and those in italics refer to grey literature sources. CDC= United States Centers for Disease Control and Prevention; MPX = monkeypox.
